# Assessment of an AI-based triage and notification system in detecting clinical signs of retinal diseases

**DOI:** 10.1101/2022.03.04.22270834

**Authors:** Manisha Agarwal, Ankita Shrivastav, Vikram Koundanya, Tanya Jain, Prashant Katre, Shibjash Dutt, Ritobroto Maitra

**Author notes:** Corresponding Author: Dr Ankita Shrivastav, Address : Radical Health-Tech Private Limited, 6, Babur, Old Fort, Saket, Saket District Centre, New Delhi – 110017. Funding Information - None. Commercial Relationship Disclosures - None.

## Abstract

**Purpose:** This study evaluates the performance of an artificial intelligence based triage and notification system that analyzes fundus photographs for nine signs: cotton wool spots, dot & blot hemorrhages, drusens, flame shaped hemorrhages, glaucomatous disc, hard exudates, retinal neovascularization, preretinal hemorrhage and vascular tortuosity. These signs may be present in multiple retinal diseases.

**Methods:** In a blinded and adjudicated study, a set of 3484 photographs of unique eyes from 3305 patients, from 15 fundus cameras, were graded by retina specialists, and the results compared with an AI-based system.

**Results:** The AI performed at a mean sensitivity of 90.19% and a mean specificity of 88.38% across all signs. The best performance was in detecting glaucomatous disc with a sensitivity of 94.65% and a specificity of 95.36%. The worst performance for sensitivity was for detecting vascular tortuosity at 85.06% and that for specificity was for detecting drusens 85.21%.

**Conclusion:** The AI-based system performs at acceptable sensitivity and specificity levels in comparison to retina specialists in a large sample pooled across 15 fundus cameras for 9 different clinical signs.

## INTRODUCTION

Artificial intelligence (AI) has the potential to positively impact health outcomes and patient care. Ophthalmology, in particular, has garnered significant attention from AI developers and this can help positively address the increasing need for ophthalmic care in the world.^[1]^ With the rising number of cases of diabetes and hypertension, as well as an increasingly aging population, the need for ophthalmic intervention in these diseases is also on the rise. In Sub-Saharan Africa there are 2.5 ophthalmologists per million population to 11.9 - 13 ophthalmologists per million in India,^[2,3]^ 46.4 and 54.7 ophthalmologists per million in the UK and the USA respectively.^[2]^ Globally there are 31.7 ophthalmologists per million population,^[2]^ which is insufficient to meet the growing demand for ophthalmic services.

Retinal complications due to diabetes and hypertension mainly produce vascular changes whereas glaucoma produces visible changes of the retinal nerve fibre layer (RNFL) and the optic nerve head (ONH). Age-related macular degeneration results in anatomical changes at the posterior pole. These diseases may not produce perceivable symptoms until the disease has reached an advanced stage. There are well-defined protocols for ophthalmic reviews and follow-ups^[4,5]^ however, adherence to these protocols are not optimal.^[6]^ Thompson et al,^[7]^ in their review on barriers in adhering to appointments, address important issues such as lack of awareness and inability to get off work. However, delays in follow-up can have adverse effects as a study from India has shown that a median time delay in meeting follow-up schedule by 5 months can convert non-vision threatening DR to vision threatening DR (vtDR).^[8]^

Artificial intelligence based devices have the capacity to scale rapidly in a very short period of time, saving the time and effort of clinicians in screening every patient. Timely diagnosis and consequentreatment measures may improve health outcomes. The current United States Food and Drug Administration (FDA) approved AI devices such as EyeArt^[9]^ and IDx-DR^[10]^ are specifically designed to diagnose more than mild DR (mtmDR) and vtDR in a pre-diagnosed diabetic population. Disease-specific algorithms will ignore signs of diseases that they have not been trained for. However, Ogunyemi et al^[11]^ have shown that 27.25% of known diabetics required a referral for diseases other than diabetic retinopathy in a specifically diabetic population. Our study evaluates the performance of RadicalEye Triage and Notification (RETN), a proprietary AI solution, in detecting clinical signs of the retina from macula-centric central fundus photographs, irrespective of the retinal disease.

Unlike IDx-DR or EyeArt, the AI device under study, RETN, is able to diagnose multiple retinal signs from a single image. The retinal signs detected by RETN are cotton wool spots, dot & blot hemorrhages, drusens, flame shaped hemorrhages, glaucomatous disc, hard exudates, retinal neovascularization (neovascularization of the disc (NVD) or elsewhere (NVE), preretinal hemorrhage and vascular tortuosity (defined in Supplementary Information table 1). A combination of these signs can help diagnose diseases such as DR (non-proliferative and proliferative), glaucoma, hypertensive retinopathy, dry AMD and many more (non-exhaustive but extensive list is provided as Supplementary Information table 2). The presence of cotton wool spots with dot and blot hemorrhages and hard exudates can indicate diabetic retinopathy whereas cotton wool spots with flame hemorrhages and vascular tortuosity are more likely to be correlated with hypertensive retinopathy. Unlike its predecessors, the use of a multi-output algorithm enables screening for multiple potential conditions instead of a single disease diagnosis. All results can be clinically correlated for further action as determined by a healthcare provider.

**Table 1.**
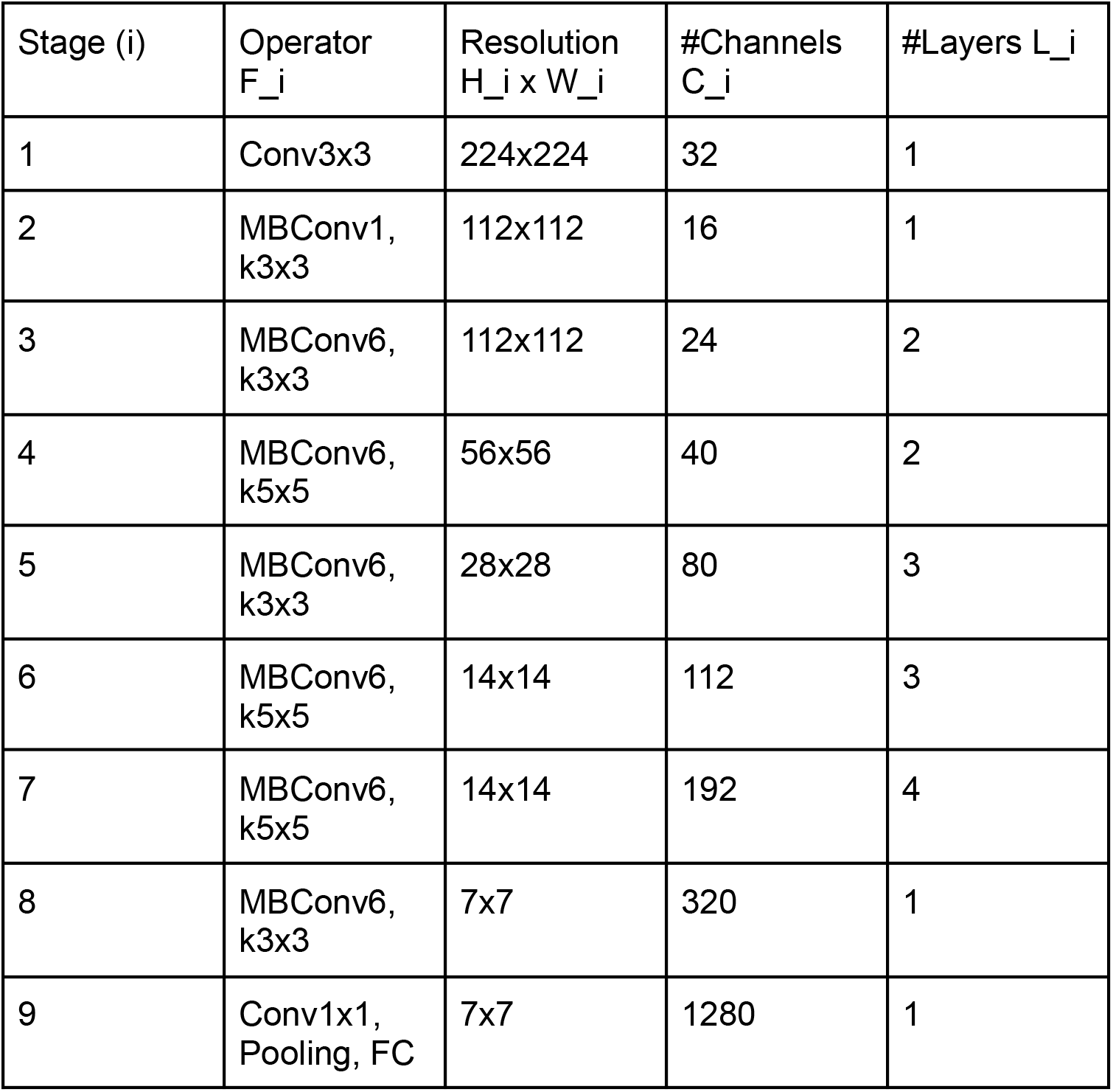
EfficientNet B0 Architecture Specification.

| Stage (i) | Operator<br>F<sub>i</sub> | Resolution<br>H<sub>i</sub> x W<sub>i</sub> | #Channels<br>C<sub>i</sub> | #Layers L<sub>i</sub> |
| --- | --- | --- | --- | --- |
| 1 | Conv3x3 | 224x224 | 32 | 1 |
| 2 | MBConv1,<br>k3x3 | 112x112 | 16 | 1 |
| 3 | MBConv6,<br>k3x3 | 112x112 | 24 | 2 |
| 4 | MBConv6,<br>k5x5 | 56x56 | 40 | 2 |
| 5 | MBConv6,<br>k3x3 | 28x28 | 80 | 3 |
| 6 | MBConv6,<br>k5x5 | 14x14 | 112 | 3 |
| 7 | MBConv6,<br>k5x5 | 14x14 | 192 | 4 |
| 8 | MBConv6,<br>k3x3 | 7x7 | 320 | 1 |
| 9 | Conv1x1,<br>Pooling, FC | 7x7 | 1280 | 1 |

**Table 2.** Sensitivity and specificity metrics for detection of each clinical sign using RETN.

| **Clinical Sign** | **Sensitivity (95% CI)** | **Specificity (95% CI)** | **Number of Positive Samples** |
| --- | --- | --- | --- |
| Drusens | 91.04% (88.03% - 94.05%) | 85.21% (83.97% - 86.46%) | 346 |
| Preretinal Hemorrhages | 89.01% (84.57% - 93.44%) | 90.59% (89.59% - 91.58%) | 191 |
| Dot and Blot Hemorrhages | 92.98% (91.42% - 94.55%) | 85.23% (83.83% - 86.63%) | 1026 |
| Cotton Wool Spots | 91.05% (88.58% - 93.52%) | 89.23% (88.11% - 90.34%) | 514 |
| Hard Exudates | 90.06% (88.28% - 91.85%) | 89.28% (88.05% - 90.52%) | 1077 |
| Flame Shaped Hemorrhages | 88.03% (86.01% - 90.05%) | 86.22% (84.87% - 87.58%) | 994 |
| Glaucomatous Disc | 94.65% (92.65% - 96.65%) | 95.36% (94.61% - 96.12%) | 486 |
| Neovascularization | 89.81% (86.52% - 93.11%) | 87.44% (86.28% - 88.59%) | 324 |
| Vascular Tortuosity | 85.06% (80.73% - 89.38%) | 86.84% (85.68% - 88.01%) | 261 |

There is a need for solutions that can triage through the population at-risk of developing visual morbidities and provide them early treatment to prevent permanent vision loss. By automating the screening process using AI, the existing health care professionals can concentrate on providing more complex eye care services. Populations who are unable to access healthcare services due to various socioeconomic barriers can avail the deeper reach of technology and seek hospital visits only when deemed necessary. Our solution can also be used to shift the focus of screening programs from opportunistic screening to a more systematic approach. RETN may also find use in the patient selection process in large clinical trials, enabling recruitment of patients who meet the inclusion criteria.

The study was conducted in accordance with the tenets of the Declaration of Helsinki. Requisite institutional review board approval was obtained from Dr Shroff’s Charity Eye Hospital, New Delhi, India, a tertiary eye care institute.

## METHODS

### Model Development

Deep learning (DL) is a subspace of machine learning (ML) which uses deep neural networks to perform various tasks. This process of training neural networks requires a large amount of labelled data since photographs for a specific disease may look different across patients, across two eyes of the same patient, or even photographs of the same eye on different fundus cameras (due to factors such as field of view, contrast and sensor of the cameras). The system employs a convolutional neural network architecture similar to EfficientNet (B6 variant),^[12]^ which represents a state-of-the-art in image classification and related tasks.

A brief specification of the EfficientNet architecture (B0 variant) is given in Table 1. We elaborate on some of the differentiating features of the EfficientNet architecture.

- MBConv: This is the Mobile Inverted Residual Bottleneck block of convolution layers, and its primary role here is to be a more efficient alternative to the typical Residual blocks popularized by ResNet. MBConv internally uses 1×1 convolutions and depthwise convolutions which also leads to lesser parameter usage.
- Swish: Contrary to most CNNs which employ the ReLU non-linearity, EfficientNet uses the Swish nonlinear function, where swish(x) = x / (1 + exp(-x * beta). Swish has been empirically found to perform better than the standard ReLU non-linearity, possibly by ameliorating vanishing gradients.
- Scaling: From the architecture of the B0 variant given in Table 1, several “scaled” variants (B1 through B7) may be generated, by systematically increasing the following architectural parameters:
  - Depth: The number of layers in each stage
  - Width: The number of channels
  - Resolution: The internal resolution (output sizes) of each layer
- EfficientNet uses a form of architecture search where each variant B0 - B7 is assigned values for the above parameters, such that each variant is more “powerful” (in the sense of predictive power) than the last, at the cost of increased parameter usage and computational requirements.
- The model was initialized from pre-trained weights that had been obtained via training on ImageNet. The weights of the output layer were randomly initialized with Xavier initialization.

#### 1. Training Data Preparation

A set of about 100,000 colour fundus photographs were curated for training a deep neural network based classifier. The training dataset is an amalgamated proprietary dataset of images that was assembled from various care centres across India by Radical. This dataset consists of fundus photographs taken from various cameras and settings and are balanced using augmentation for various contextual metadata (such as field of view, age, sex or camera) as well as clinical sign prevalence, but all algorithm training processes are blinded to such details. When such balancing is not possible using simply sampling from a dataset, augmentation techniques are used. The principle of training on a blinded basis is done to ensure that inadvertent errors of bias do not creep into the dataset, and because no output of the device is designed to take into consideration any of the image metadata. The network is supposed to work solely on pixel data with a set of labels, and it is trained correspondingly.

Further details of the training dataset are provided in the supplementary information for clinical data..

#### 2. Labelling Criteria, Quality Control

Fundus images were annotated by graders on a web-based tool. Graders were asked to identify the clinical signs exhibited in the fundus image. In addition, they were also provided an option to signal the presence of imaging artefacts, such as camera flash, dust spots, lens flare and others. Further, the graders also had the ability to indicate if the image had no diagnostic value, based on the visibility of the retinal anatomical landmarks of the disc, macula and major arcade vessels. Thus, for each labelled image, grading produced a corresponding bit-string indicating the presence or absence of various signs, another one corresponding to imaging artefacts, and a flag indicating undiagnosable images. Images that were marked as undiagnosable were excluded from the dataset and were not utilized in subsequent processing and training.

For clinically validating the performance of the trained model, we additionally collected a validation set of images, and followed a rigorous process in labelling these images. The details and the subsequent statistical analysis of the model performance are described later.

#### 3. Training Data Selection : Image Pre-processing and Augmentations

Images were close-cropped, removing any borders and padding. They were then resized, preserving aspect-ratio such that the longer side was 384 pixels long. To aid regularization during neural network training, and to produce a more robust model,extensive augmentations were applied. Augmentations were applied probabilistically to each batch of images fed into the neural network after choosing control parameters from predefined ranges. Both geometric transformations (rotations, translations, random cropping and shearing) as well as color transformations (altering the hues of the image slightly, converting a fraction of images to grayscale, varying intensity/lightness, color channel shuffling and dropping) were used. Augmentations that artificially simulate camera sensor noise were also used. Additionally, the MixUp^[13]^ technique was used at training time. MixUp has been found to promote model generalization and lessen the impact of incorrect labels.

#### 4. Training Details

Prior to training, training/test splits were first generated by stratifying across patients, in order to ensure that multiple images from the same patient do not end up across splits and poison the resulting metrics. This stratification process ensured that the training and validation data subsets were completely disjoint i.e. no images were present in more than one set, nor did images from a given patient appear in multiple sets. To predict the clinical signs in questions, an EfficientNet based model architecture was used. The underlying deep-learning based algorithm is designed to be used alongside a standard (non-wide field) fundus camera, but it can accept fundus images that have been directly uploaded as well. Macula-centered fundus images in formats DCM, JPEG, PNG, BMP, TIFF or DICOM act as inputs to the algorithm which detects a suspected retinal clinical sign. The device does not produce the location of such a sign. Since any of the 9 clinical signs in question may occur independently of each other, the problem was modelled as a multi-label classification task, wherein a single model simultaneously predicts each sign.

There are certain advantages to using a single large multi-label model over 9 smaller distinct binary output models. The former may be taken as an instance of Multi-Task Learning, which forces the model to learn feature representations that are generic and help contribute to all the different outputs. In practice, this leads to better generalization, and better predictive performance. Training 9 distinct models also transforms the training objective from a multi-label loss into 9 one-vs-rest binary losses trained separately. The former is more useful, since it allows the model to take into consideration factors like sign co-occurrence and comorbidity.

The usual classification head of the EfficientNet architecture was replaced with a so-called Linear layer with 9 outputs. The entire model was trained with the Adam optimizer, with weight decay and cosine learning rate scheduling and early stopping for a total of 25 epochs, until the test loss did not decrease any further. Initial values for hyperparameters such as learning rate were chosen using random search.

### Algorithm Output

As described above, the model’s output is from a so-called Linear (or Dense) layer that produces output logits as a tensor of shape (N, 9) where N is the batch-size (number of images in a batch) and 9 corresponds to the 9 clinical signs in question.

The logits are then scaled via the sigmoid operation to lie in the range [0, 1].

This activation score is then transformed into a binary decision output (0 or 1) by using thresholds for each clinical sign, where the threshold for each sign was determined from a small held-out “development set” by maximizing Youden’s J index; an approach that equally penalizes Type - I and Type - II errors.

Thus, for each input image, corresponding to each of the signs we obtain 0 or a 1 indicating the absence or presence of that sign as judged by the model.

### Image Grading for Validation

Image grading for validation used the same workflow software as image grading for training, but the protocol followed was different. All photographs were first anonymised of any associated metadata, such as camera or patient information, prior to being labelled by the retina specialists. Each photograph was then labelled independently by two retina specialists. Each specialist labelled the clinical signs that were present in the photograph as per internationally accepted standards and definitions of those clinical signs.^[14-16]^ In case both specialists deemed a photograph to be of insufficient diagnostic quality, the photograph was removed from consideration. One photograph often contained more than one clinical sign depending on the underlying disease or diseases and their progression.

In case there was any difference in the labels of two retina specialists, the photograph was further labelled by a third, senior specialist, as an adjudicator. “Any” difference implies that the two sets of labels did not match each other exactly. For example, one reader could indicate the presence of 3 clinical signs in the photograph. Unless the other reader also indicated those exact 3 labels, the photograph was forwarded for adjudication. The two readers and the adjudicators were blind to any labels assigned to the photograph previously by another reader, or the AI system. The labels associated with the photograph were determined to be either the consensus of the two initial readers in the case of an exact match, or the final adjudicated label. There were 5 readers and 1 adjudicator in this study.

Parallel to this, the algorithm also produced the set of labels for each photograph. The algorithm outputs a binary vector with 9 elements for each input image. These correspond to the nine clinical signs detected by the model. A value of 0 indicates a lack of the corresponding sign in the image, a value of 1 indicates that the sign has been detected in the input. These outputs were then compared to the gold standard validation described above, for each individual clinical sign, and metrics were hence derived.

### Sample size

A large retrospective set of 28308 fundus photographs was initially considered to be the pool of population samples available for study. These samples were initially shuffled randomly before being graded sequentially by human expert readers, i.e, vitreoretinal specialists. The following standard assumptions were made in calculating the sample size required for statistical analysis of sensitivity and specificity for each clinical sign - expected sensitivity: 90%, expected specificity: 80%, prevalence: 20%, confidence level: 95%, dropout rate: 10%. This produced a minimum sample size for sensitivity as 173 samples, and that for specificity at 77 samples. So, a minimum of 77 negative samples were required for each sign, and a minimum of 173 positive samples were required for each sign. Considering dropout, the minimum sample size was fixed at 191 samples. The requisite theoretical framework and formulae used to derive these are available in the work of Buderer.^[17]^ This sample size was verified against other methods of estimating sample sizes such as binomial estimation, and were found to be similar or exact, thereby providing additional confidence regarding the size of the sample. As a measure of literature review, the sample sizes for triage and notification devices approved by the US FDA were studied and were found to be similar as well. It was safe to assume that the number of minimum samples required would be achieved throughout the study without specific enrichment strategies since a sample without any of the 9 clinical signs would serve as a common pool of negative samples for all of the 9 clinical signs. This assumption was found to be correct in the course of the analysis.

Based on this requirement, samples were enrolled into the validation set until this sample size of 191 positive samples was reached for all 9 clinical signs under consideration. In the course of the study, the last set to reach this threshold were positive samples for preretinal hemorrhages, and at that point further enrolment of samples into the study was stopped, and the final sample set for the study was considered final and frozen.

In creating the original gold standard benchmark data set for evaluation, a total of 55962 sets of labels from 28308 eyes were created across 14718 patients by the readers. Photographs were then eliminated if they only contained signs outside the scope of the study, and undiagnosable photographs were removed. Further, photographs of the same eye were removed, leaving a single image per eye at random. After this, the final validation set of 3484 photographs of 3484 unique eyes from 3305 patients was frozen.

### Statistical Analysis

There were two primary metrics of interest in the study; namely, the sensitivity and specificity of the algorithm in detecting each of the 9 clinical signs. 95% confidence intervals were calculated as standard binomial intervals. All calculations were done using Python scripts using numpy and scipy. Table 2 enumerates the results of the performance testing. In addition to the primary metrics of sensitivity and specificity, we also wished to report the variability in the performance of the algorithm with respect to age, sex, camera and field of view. This was done to evaluate statistically whether the error rates of the algorithm (false positives and false negatives) were independent of the above factors, or were different for different characteristics. The independence of the algorithm to the above factors is not the primary endpoint. Nonetheless, we report some signals and leave further investigation to a future work.

In calculating these results, chi-square tests of independence were used to establish whether or not the categorical variables of the study showed any dependence on the outcomes. The null hypothesis was that the rate of false positives and false negatives for different categories would be independent of the category. A p-value of less than 0·05 would indicate that the null hypothesis needed to be rejected, ie, the rates of error in the algorithm are dependent on the covariate. A p-value of more than 0·05 would indicate that the null hypothesis was true, ie, no effect of the categories of the variable under investigation, such as different types of camera models or different sexes were seen on the rates of error of the algorithm. If a certain variate had very few or insufficient samples, they were removed. (Table 3) (detailed description of this data is available as Supplementary Information of clinical data for each individual clinical sign).

**Table 3.**
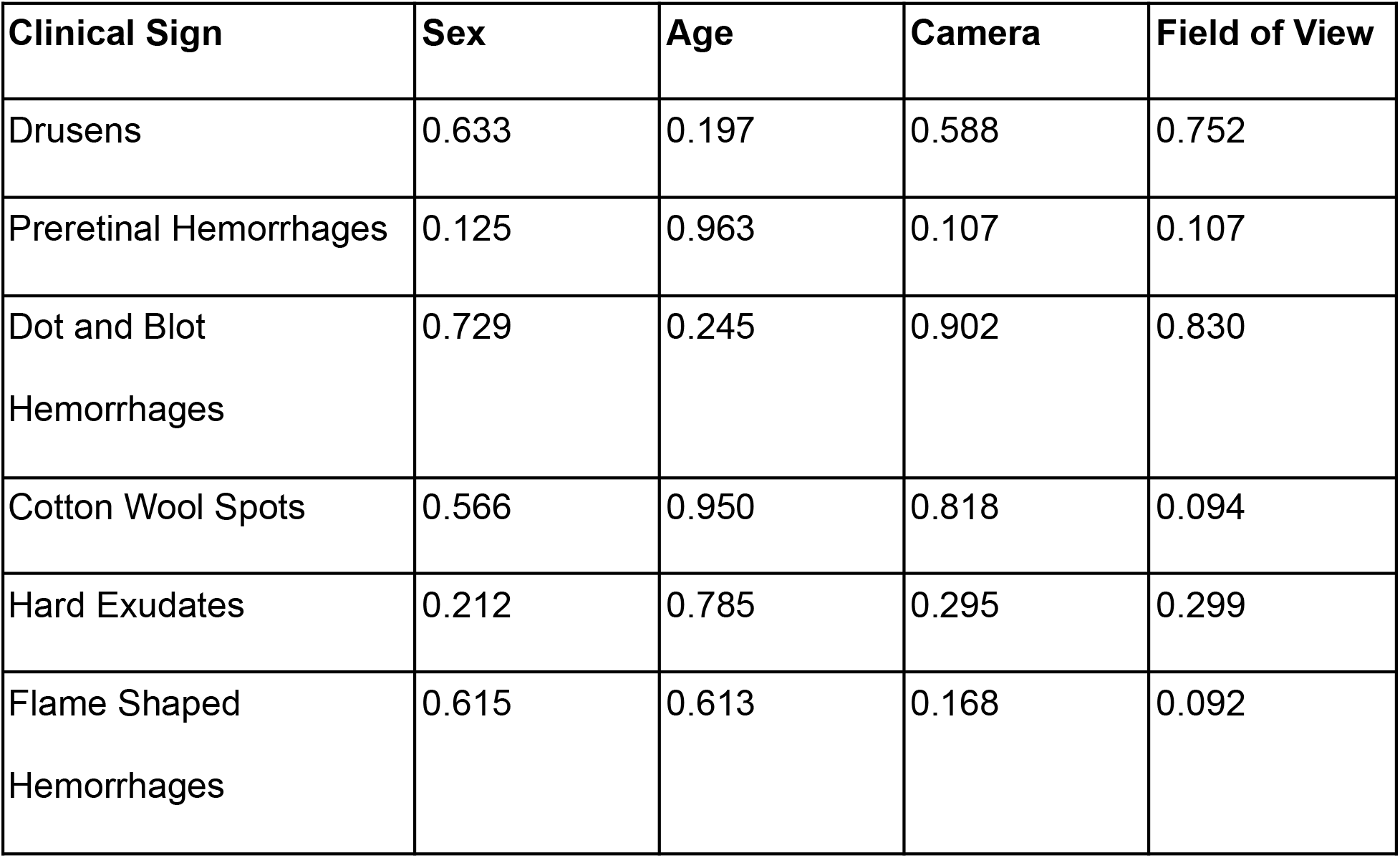
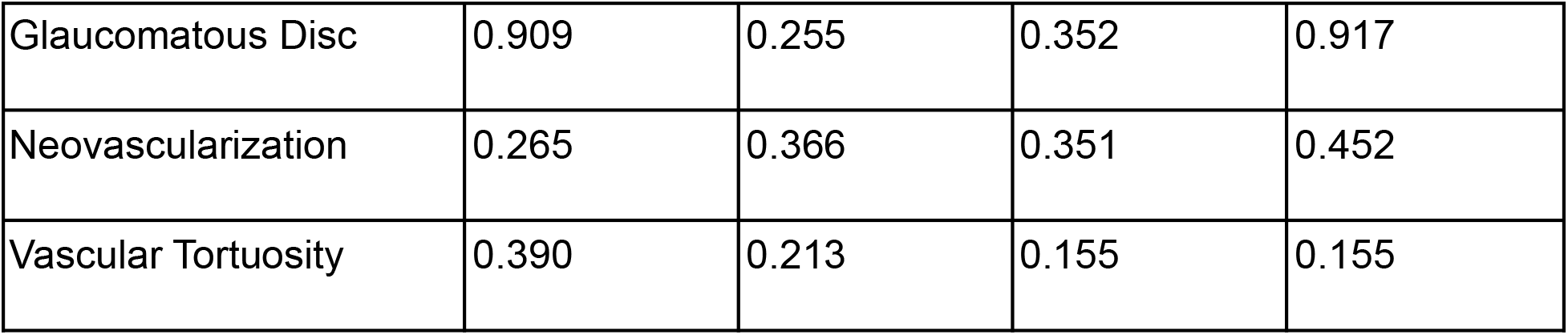
Effects of sex, age, camera and field of view on the error rates of the deep learning algorithm. All p-values above 0.05 indicate no statistically significant effect.

## RESULTS

### Description of Data

RadicalEye Triage and Notification was validated on a set of 3484 retrospective macula-centred fundus photographs acquired through various fundus cameras at multiple clinical sites at tertiary and secondary eye care facilities in India. All data was captured in regular clinical workflow and reflects a real-world setting of an ophthalmic photographer or optometrist capturing the photograph on various cameras.

A total of 15 different types of fundus cameras were used in the study to study any statistically significant variation in the errors in the algorithm. Further, they had 6 different fields of view (the same camera could also be operated on different fields of view). Each clinical feature had photographs from at least 2, at most 7 and on average 4.4 cameras (refer to Supplementary Information of clinical data for more information). Further, since the algorithm does not diagnose a specific disease but looks at specific clinical signs, such as hemorrhages and exudates, they are racially invariant. The prevalence of a disease may be influenced by race, ethnicity or other clinical factors, but the underlying clinical signs that are related to a disease do not vary.

Undiagnosable photographs refer to photographs where the following retinal landmarks are not visible: the optic disc, the macula and the major arcade vessels. They do not refer to photographs where these may be occluded by a clinical feature, for example, a preretinal hemorrhage. No two images belong to the same eye.

1080 photographs did not have any of the signs under consideration. 2404 photographs had at least one sign under consideration, with a maximum of 6 signs in a single image and an average of 2.17 signs per image.

Of the 3484 images considered in total, the number of positive samples for each clinical sign under consideration as per the gold standard data set created as in Table 4. The two sexes in the dataset were male (64.2%) and female (35.8%). The age categories in the dataset were: 18 to 29 years old, 30 to 39 years old, 40 to 49 years old, 50 to 59 years old, 60 to 69 years old, and 70 years or above. The mean age was 50.19 years (SD: 28.06 years).

**Table 4.** Number of positive samples for each clinical sign under consideration.

|  |  |
| --- | --- |
| Drusens | 346 |
| Preretinal Hemorrhages | 191 |
| Dot and Blot Hemorrhages | 1026 |
| Cotton Wool Spots | 514 |
| Hard Exudates | 1077 |
| Flame Shaped Hemorrhages | 994 |
| Glaucomatous Disc | 486 |
| Neovascularization | 324 |
| Vascular Tortuosity | 261 |

The different fields of view in the dataset were (degrees): 30, 35, 40, 45, 50 and 60. The different fundus cameras in the dataset were as follows: Canon CR-5, Canon CR-1, Canon CF-60UVi, Film Camera Scan (Scanned photographs from non-digital, film-based fundus cameras), Forus 3Nethra Classic, Kova VX-10, Nidek AFC-210, Nidek NM-200-D, Topcon TRC-NW400, Topcon 50DX, Topcon TRV-50, Topcon TRC-NW6, Zeiss FF450 Plus, Zeiss Visuscout and Zeiss Visucam 500.

### Performance Characteristics: Primary Metrics

Since this system analyses the photograph for 9 different signs, we also present a set of macro, averaged metrics for ease of understanding and communication in Table 5. The overall mean was 90.19% for sensitivity and 88.38% for specificity.

**Table 5.** Macro metrics of sensitivity and specificity for nine clinical signs detected by RETN.

| **Macro Performance Metric** | **Sensitivity** | **Specificity** |
| --- | --- | --- |
| Best | 94.65% | 95.36% |
| Worst | 85.06% | 85.21% |
| Arithmetic Mean | 90.19% | 88.38% |
| Median | 90.06% | 87.44% |

### Performance Characteristics: Secondary Metrics

The results of the algorithm were found to be independent of age, sex, camera and field of view. Had it been dependent, it would not be possible to indicate the system for use irrespective of the above factors. It should be noted that the same camera can sometimes be operated at different fields of view depending on the operator and the manufacturer or mode of use, and hence field of view is considered a separate measure from the camera itself. The data considered in this study contains no wide-field fundus camera or ultra-wide field fundus camera and hence the algorithm should be considered untested on wide field fundus camera photographs.

A further performance characteristic that was measured was the average time for inference. The time was measured on inference over all samples in the validation set on Amazon Web Services (AWS) cloud servers. They were measured on p3.2xlarge instances on AWS, in batch mode with a batch size of 32. As an intended cloud-based multi-tenant solution, this represents the intended use scenario for the algorithm. GPU evaluations were carried out on an NVidia Tesla V100 GPU. CPU evaluations were carried out on an Intel CPU (Xeon E5-2686v4 (2.30 GHz) with 8 Cores). For CPU evaluations, inference was parallelised across all cores. The mean time taken on GPU-accelerated inference was 2.74 milliseconds (ms) per image (SD: 2.64 ms) and the mean time taken on CPU inference was 222.48 ms per image (SD: 10.76 ms). For most use cases, an optimised and CPU-accelerated inference would suffice (with the capacity to analyse 2 patients per second per server considering one photograph per eye), but for critical and near-instantaneous results, GPU-acceleration for inference could be used (with the capacity to analyse 182 patients per second per server considering one photograph per eye).

## DISCUSSION

### Clinical Applicability

The core algorithmic principle upon which the system runs is the analysis of a retinal fundus photograph for the presence of one or more of the nine clinical signs mentioned above. These clinical signs individually and in combination are suggestive of various ophthalmic diseases such as DR, AMD, hypertensive retinopathy, glaucoma, retinal vein or artery occlusions, and vasculitis among many others. The model is useful in scenarios where retina specialists are not available as it can flag the abnormal signs and suggest the need for a more detailed evaluation, both of the eye and the systemic diseases that the retinal pathologies may indicate. The model helps to detect the nine retinal signs and may be used in various settings such as screening camps and diabetic clinics. However it does not preclude the need for a comprehensive examination. It may also find application in ophthalmology hospitals to direct patients to appropriate specialty clinics as per the findings of the scan. For example, a fundus photograph with a glaucomatous disc merits to be seen by a glaucoma specialist whereas a photograph with retinal hemorrhages is more suited for a referral to a retina specialist.

Making the model part of screening programs makes it possible for a much larger population to be screened for retinal diseases without the burden of availability of specialists for the same. This in turn leads to an early diagnosis. As most retinal diseases are vision threatening if not treated timely, an early identification allows significant improvement in the patient’s quality of life and reduces financial burden of treatment.

As opposed to screening programs which can be conducted at large scales using the RETN model, programs conducted with physical eye examination may have limited capacity and long waiting time for appointments may lead to a delayed diagnosis leading to advanced stages of the disease, resulting in frequent complications and requiring more expensive interventions.

For glaucoma, the cost of treatment increases from $455 in the earliest stage doubling to $969 in the advanced^[18]^ stages. Similarly, it has been studied that even a 6 month delay in treatment of vein occlusion can lead to a loss to 9 to 11 ETDRS Letters.^[19]^ Specifically for AMD, early treatment at non-neovascular stages can be effective in halting the progression of disease. 84% of individuals with AMD in the US are unaware of their disease.^[20]^ It is especially important since drusens usually present asymptomatically, and once the disease progresses to wet or neovascular AMD, untreated patients can progress rapidly. One line of visual acuity loss is reported as early as 3 months.^[21]^ Once AMD reaches the stage where CNV has developed, progression of CNV can be rapid, with immature vessels reaching a maturation state within 10–14 days,^[21]^ and patients may remain asymptomatic during this growth. Using RETN, a more active and relatively more comprehensive screening can be envisioned rather than patients seeking care when they experience symptoms of disease. However, we acknowledge that detection of some signs such as glaucomatous disc subjects itself to variability even between two glaucoma specialists and hence clinical correlation is advised on clinical signs triaged on RETN.

Retina is a mirror of systemic diseases. For example, flame shaped hemorrhages on a retinal image can be related to retinal vein occlusion (RVO), which in turn may be manifestation of hypertension or cardiovascular disease (CVD).^[22]^

The nine retinal signs identified by RETN, may be common to several retinal and systemic diseases. For example, systemic lupus erythematosus (SLE) presents with signs similar to DR such as cotton wool spots and retinal hemorrhages.^[23]^ Specific types of anemias may also mimic DR. For example, sickle cell retinopathy can simulate a DR fundus in both its proliferative and pre-proliferative stages.^[24]^ Radiation retinopathy^[25]^ can also present with intraretinal hemorrhages.

Cotton wool spots develop in areas of retinal hypoperfusion and can be seen in diseases like DR, hypertensive retinopathy, and also infectious diseases such as those caused by the human immunodeficiency virus (HIV).^[26]^ The presence of drusens in younger populations may point towards underlying inflammatory diseases.^[27-29]^

The output from RETN may help in triaging the patients based on their retinal signs identified thereby helping them seek early medical advice. The nine clinical signs detected by RETN are among some of the common signs seen on a fundus photograph and represent most major retinal diseases (refer to Supplementary Information table 2 for a complete list). The algorithm is agnostic to use cases. For instance, the model may be used to triage and prioritize cases in a patient list at an eyecare institution. It may also be used in non-ophthalmic setups such as a diabetic clinic or optical store for referral to ophthalmologists based on the generated report. Each of these settings will present a population with different prevalence rates for the various signs. For instance, the population at a diabetic clinic will self-select for a higher prevalence of hard exudates and dot and blot hemorrhages. Thus, certain statistical measures, such as positive predictive value (PPV) and negative predictive value (NPV), which are linked to prevalence, will vary. To take this into account, we have included the expected statistical performance across a range of prevalences as a table 3 in the supplementary information.

### Comparison with State of the Art

In our study, we found that among all clinical signs, the worst sensitivity of RETN was 85.06% for vascular tortuosity and that of specificity was 85.21% for drusens. The best sensitivity and specificity was 94.65% and 95.36% respectively for glaucomatous disc.

On review of literature, we found that these metrics in isolation are comparable to the ones previously reported by various authors such as Gulshan et al,^[30]^ who reported a sensitivity of 97.5% and a specificity of 93.4% (at an operating point for high sensitivity for detecting referable DR), Rajalakshmi et al^[31]^ reported a sensitivity of 95.8% and 80.2% specificity (for detecting DR), Abramoff et al^[10]^ have reported a sensitivity of 87.2% and a specificity of 90.7% (which was also used by the FDA in approving their algorithm for diagnostic use for detecting DR), Burlina et al^[32]^ reported an accuracy between 88.4% and 91.6% for detecting AMD, Olvera-Barrios et al^[9]^ reported sensitivity of any retinopathy ranging from 92.26% to 92.27%. However, all these algorithms previously reported were focussed on detecting specific diseases, whereas this study focuses on simultaneous detection of nine retinal signs.

It is useful to compare the current system under evaluation with previously published work. On comparing RETN with the existing devices such as IDx-DR and EyeArt, the major advantage is that this device flags the various clinical signs irrespective of the underlying cause and thereby not restricting itself to diagnosing a particular pathology such as DR. Cotter et al emphasized that even in images unreadable for DR, other important pathology may be captured and hence it is important for screening tools to be able to diagnose other treatable eye diseases in addition to DR.^[33]^

IDx-DR is designed to detect one specific disease, more than mild diabetic retinopathy in pre-diagnosed populations. RETN is a triage and notification device designed to notify the user for the presence of medical signs, not one specific disease. Hence, the device outputs are entirely different. IDx-DR is indicated for use on a pre-diagnosed diabetic population. This device is designed to be used for any patient who the ordering provider believes has the potential for having any of the nine clinical signs identified by the algorithm, regardless of whether the patient has current symptoms suggestive of disease. In other words, there is no preset diagnostic or other restriction on the population for which RETN is designed to be used. Hence, the populations for which the devices are designed to be used are different (RETN’s population is a superset of IDx-DR’s population). IDx-DR is indicated for use only with the Topcon NW-400 Fundus Camera. RETN’s performance is invariant over multiple fundus cameras, and hence is not restricted to be used with a specific fundus camera.

IDx-DR is labelled for use autonomously. RETN however is not envisioned to be a standalone diagnostic device, and the results must be correlated clinically. It is not designed to be a diagnostic device with the capability to definitively rule out or in any of the signs listed above. Hence, it is appropriate for use by a clinician at their discretion. The clinical judgement of the ordering physician determines the subject’s diagnosis, treatment and other clinical activity.

A feature of RETN is the use of a multi-label classifier. A binary classifier typically produces an output which classifies an image into one of two classes (such as, presence or absence of diabetic retinopathy). Previous work on analyzing multiple signs from a single fundus photograph have been demonstrated in the work of Son et al.^[34]^ In that work, the authors evaluated a system with the ability to detect 12 clinical signs. However, they employed 12 different binary classifiers in each of these clinical signs. It was noted in editorial commentary of their work^[35]^ that a future direction of work could be to produce ensemble models that aggregated all 12 binary classifiers. The current system under consideration addresses that issue, by using multi-label classification on a single algorithm, rather than creating nine different models. This has the potential to be much easier to maintain one single algorithm rather than multiple ones. Moreover, deep learning algorithms are essentially hierarchical in nature. The lower layers of these neural networks learn low-level features, which can be shared among many different higher level tasks (of learning to recognise multiple clinical signs) without duplication and re-training. It is generally known that such types of learning algorithms (generically known as multi-task learning algorithms) provide advantages in performance when the underlying distribution of data is similar. Son et al also mention vascular abnormalities as one of the detected abnormalities but do not subclassify them. In contrast, RETN is capable of diagnosing vascular tortuosity and neovascularization as two specific instances of vascular anomalies in the retina.

### Limitations and Future Directions

The number of clinical signs that exist in the retina is substantially larger than what has now been considered in the scope of this study. A definite future direction would be to evaluate the efficacy of the system in being able to detect more clinical signs than are currently indicated. General principles of deep learning provide an intuition that the single-model approach that is currently used may provide a stronger basis for adding more signs and an easier pathway towards improvements rather than training a new model from the beginning for every new sign. A primary metric of interest in future studies may be long-term and prospective settings to evaluate the time and costs saved in clinical evaluation or referrals by triaging patients, as well as measurable benefit to patient health. Another direction of work would be to analyze the effectiveness of the same or a similar system on ultra-wide field camera photographs. These photographs are substantially different from any camera photograph with less than 60 degrees field of view. Future AI models may be used to predict the natural history of each clinical sign and disease process, in turn substituting the control arm by virtual controls. This may reduce the burden of recruitment significantly. The efficacy of this system has not been evaluated on non-macula centered fundus photographs. Future studies should also include conducting the trial on different races and in different geographies to make the results more inclusive.

## Supporting information

Supplementary clinical data

## Data Availability

All data produced in the present work are contained in the manuscript

## Acknowledgements

The authors would like to acknowledge the valuable feedback, suggestions and support received from Dr Christopher Jankosky, MD, WorkCare Inc., and Don K. Dennis, Carnegie Mellon University, in early reviews of the draft. The authors would like to acknowledge the support provided by members of Radical Health-Tech Private Limited (Sonu Jaglan, Shubham Rateria and Prashant Baghel), members of WorkCare, Inc. (Dr Peter Greaney), members of Amazon Web Services, Inc. (Kapil Bansal, Mayank Raja), the members of Dr Shroff’s Charity Eye Hospital (Dr Shalini Singh, Dr Rahul Mayor, Dr Chanda Gupta, Dr Shalin Shah, Brajesh Kumar), the members of Disha Eye Hospitals (Dr Debasish Bhattacharya, Dr Samar Kumar Basak, Dr Debdulal Chakraborty, Dr Soham Basak, Ranabir Bhattacharya, Utpal Sarkar and Sankha Ghosh) for their guidance and contributions.

## Notes

### Competing Interest Statement

The authors have declared no competing interest.

### Funding Statement

This study did not receive any funding

### Author Declarations

Ethics committee and institutional review board of Dr Shroff's Charity Eye Hospital, New Delhi

