## Supplementary Table 2 for "Assessment of an AI-based triage and notification system in detecting clinical signs of retinal diseases"

**Supplementary Information**

**Table**

Indicative, Non-Exhaustive, Non-Prioritised List of Differentials for Clinical Signs detected by RETN:

| **Clinical Sign** | **Age related diseases** | **Vascular diseases** | **Systemic diseases** | **Infectious diseases** | **Hereditary diseases** | **Tumors** | **Others** |
|---|---|---|---|---|---|---|---|
| **Drusen** | Age-related Macular Degeneration (AMD) | | Glomerulonephritis | | North Carolina Macular Dystrophy, Familial Dominant Drusen | Choroidal Nevus | Central Serous Chorioretinopathy (CSCR) |
| **Preretinal Hemorrhage** | | Diabetic Retinopathy (DR), Retinal Vein Occlusion (RVO), Sickle Cell Retinopathy, Retinal Artery Macroaneurysm (RAM) | Diabetes, Hypertension | | X-linked Juvenile Retinoschisis | Leukemic Retinopathy, | Terson's Syndrome, Shaken Baby Syndrome, Retinal Tear, Posterior Vitreous Detachment (PVD) |
| **Dot and Blot Hemorrhage** | | DR, Hypertensive retinopathy (HTR), RVO, Anemia, Ocular Ischemic Syndrome, RAM, Sickle cell retinopathy, Bone marrow | Diabetes, Hypertension, Sarcoidosis | Human immunodeficiency virus (HIV) retinopathy, Cytomegalovirus (CMV) Retinitis | | Leukemic retinopathy | Intraocular Gentamicin, Behcet's Disease, Blunt Trauma, Shaken Baby Syndrome, Valsalva Retinopathy, Terson's Syndrome, Purtscher's |

| | | | | | | | |
|---|---|---|---|---|---|---|---|
| | | transplantation, High altitude retinopathy | | | | | Retinopathy, Vitreomacular Traction Syndrome, Retinal Tear, Retinal Detachment, PVD, Radiation retinopathy |
| **Cotton Wool Spot** | | DR, Hypertensive Retinopathy, RVO, Anemia | Diabetes, Hypertension, Systemic Lupus Erythematosus, Behçet's Disease, Giant Cell Arteritis, Polyarteritis Nodos | HIV Retinopathy, CMV Retinopathy, Cat Scratch Disease, | | Leukemic Retinopathy | Bone Marrow Transplantation, Interferon Toxicity, Purtscher's Retinopathy, Blunt Trauma, Radiation Retinopathy |
| **Hard Exudate** | AMD | DR, Hypertensive retinopathy, RVO, Juxtafoveal Retinal Telangiectasis, RAM, Coat's disease | Diabetes, Hypertension | | | Choroidal tumours | CSCR, Chronic Retinal Detachment, Radiation Retinopathy |
| **Flame Shaped Hemorrhage** | | Hypertensive retinopathy, DR, RVO, Retinal vasculitis, Anemia, Ischemic Optic Neuropathy | Diabetes, Hypertension, Vitamin D deficiency, Sarcoidosis | Acute bacterial endocarditis | | Leukemic retinopathy | |
| **Glaucomatous disc** | | Ischemic Optic Neuropathy, Chronic | | | | | Glaucoma, Methanol Poisoning, Compressive |

| | | | | | | | |
|---|---|---|---|---|---|---|---|
| | | Retinal Vessel Occlusion | | | | | Optic Neuropathy |
| **Neovascularization** | | DR, RVO, Sickle cell Retinopathy, Retinopathy of Prematurity, Eale's disease | Diabetes, Hypertension, Sarcoidosis | | Familial Exudative Vitreoretinopathy, Norrie's Disease, Incontinentia Pigmenti | Leukemic Retinopathy | Talc Retinopathy, Pars Planitis, Chronic Retinal Detachment, Radiation Retinopathy |
| **Vascular Tortuosity** | | Hypertensive Retinopathy, Chronic anemia | Hypertension, Coronary Artery Disease, Obesity, Obstructive Sleep Apnea | | | | |
