## Supplementary Table 1 for "Assessment of an AI-based triage and notification system in detecting clinical signs of retinal diseases"

Supplementary information Table 1: Appendix to definitions of signs detected by RETN

| **Clinical Sign** | **Definition** |
| --- | --- |
| ***Drusens*** | - Discrete yellow-white sub RPE deposits - Single drusen is round typically round in shape - Drusens may coalesce to form confluent drusen or raise a drusenoid PED - Hard drusen is more well defined than soft drusen. - For the purpose of labelling, we are not distinguishing between soft/hard/reticular drusen. |
| ***Preretinal Hemorrhages*** | - Heme in the preretinal space - May often appear linearly elongated (boat shape) - Obscures the visibility of retinal vessels |
| ***Dot and Blot Hemorrhages*** | - Dark red, circular, well defined red dots that are larger than microaneurysms which appear as pinpoint red lesions. Blot hemorrhages are red lesions with ill-defined borders (ink-blot shape). - These hemorrhages do not occlude the visibility of retinal vessels. - They must be differentiated from round artefacts. Artefacts are very sharp, appear on multiple images. |
| ***Cotton Wool Spots*** | - May be seen arranged in the pattern of retinal nerve fibre layer - Yellow-white or gray-white fluffy lesions - Appear cloud-like, linear or serpentine lesions with fimbriated borders - Presents in the superficial retina - Appears elevated |
| ***Hard Exudates*** | - Yellowish deposits of lipid and protein in the intraretinal space - Irregular in shape, have sharp borders |
| ***Flame Shaped Hemorrhages*** | - Red in colour, superficial hemorrhages - Flame or feather shaped hemorrhage with indistinct borders - Spindle shaped - Often follows the arrangement of the retinal nerve fibre layer |
| ***Glaucomatous Disc*** | - Presence of one or more of the following :   - Optic disc cupping > 0.6   - Beta zone parapapillary atrophy   - RNFL Defect   - Disc hemorrhage   - NRR notching |
| ***Neovascularization*** | - Mesh of fine new vessels appearing on the surface of the retina. Includes -   - Neovascularization at the disc (NVD) - Neovascularization at or within 1 disc diabetes of the optic nerve head or   - Neovascularization elsewhere (NVE) - Neovascularization on the retina at any other location other than described above |
| ***Vascular Tortuosity*** | - Any vessel that has an abnormal number of twists and turns per unit length of the vessel when compared to the normal fundus image. - We shall refer to this convoluted / corkscrew appearance of the vessel as a local arc formation. The height of this arc, when more than 2/3rd the chord length of the arc, will be defined as vascular tortuosity. |
